# Using Twitter Data for Cohort Studies of Drug Safety in Pregnancy: A Proof-of-Concept with Beta-Blockers

**DOI:** 10.1101/2022.02.23.22271408

**Authors:** Ari Z. Klein, Karen O’Connor, Lisa D. Levine, Graciela Gonzalez-Hernandez

**Author notes:** Corresponding author: Ari Z. Klein, Blockley Hall, 4^th^ Fl., 423 Guardian Dr., Philadelphia, PA 19104, USA.

## Abstract

**Background:** Despite that medication is taken during more than 90% of pregnancies, the fetal risk for most medications is unknown, and the majority of medications have no data regarding safety in pregnancy.

**Objective:** Using beta-blockers as a proof-of-concept, the primary objective of this study was to assess the utility of Twitter data for a cohort study design—in particular, whether we could identify (1) Twitter users who have posted tweets reporting that they took a beta-blocker during pregnancy and (2) their associated pregnancy outcomes.

**Methods:** We searched for mentions of beta-blockers in 2.75 billion tweets posted by 415,690 users who announced their pregnancy on Twitter. We manually reviewed the matching tweets to first determine if the user actually took the beta-blocker mentioned in the tweet. Then, to help determine if the beta-blocker was taken during pregnancy, we used the timestamp of the tweet reporting intake and drew upon an automated natural language processing (NLP) tool that estimates the date of the user’s prenatal time period. For users who posted tweets indicating that they took or may have taken the beta-blocker during pregnancy, we drew upon additional NLP tools to help identify tweets that report their adverse pregnancy outcomes, including miscarriage, stillbirth, preterm birth, low birth weight, birth defects, and neonatal intensive care unit admission.

**Results:** We retrieved 5114 tweets, posted by 2339 users, that mention a beta-blocker, and manually identified 2332 (45.6%) tweets, posted by 1195 (51.1%) of the users, that self-report taking the beta-blocker. We were able to estimate the date of the prenatal time period for 356 pregnancies among 334 (27.9%) of these 1195 users. Among these 356 pregnancies, we identified 257 (72.2%) during which the beta-blocker was or may have been taken. We manually verified an adverse pregnancy outcome—preterm birth, neonatal intensive care unit admission, low birth weight, birth defects, or miscarriage—for 38 (14.8%) of these 257 pregnancies.

**Conclusions:** Our ability to detect pregnancy outcomes for Twitter users who posted tweets reporting that they took or may have taken a beta-blocker during pregnancy suggests that Twitter can be a complementary resource for cohort studies of drug safety in pregnancy.

## Introduction

Medication use during pregnancy has increased by 68% over three decades, and it is estimated that prescription or over-the-counter (OTC) medication is taken during more than 90% of pregnancies [1]. Despite the widespread use of medication during pregnancy, the fetal risk for most medications approved by the United States Food and Drug Administration (FDA) is unknown, and the majority of approved medications have no data regarding safety in pregnancy [2]. Therefore, additional sources of data for evaluating drug safety in pregnancy should be explored. In the United States, 42% of people aged 18-29 and 27% of people aged 30-49 use Twitter [3], and our prior work [4] used Twitter data in a case-control study that involved identifying users who reported a birth defect outcome (cases) [5] and users who did not (controls), and then searching their tweets for reports of medication exposure during pregnancy. Twitter data has not been assessed, however, for its utility in a cohort study design, which would involve identifying pregnancy outcomes for users who have reported taking medication during pregnancy. Using beta-blockers as a proof-of-concept, the primary objective of this study was to assess whether we could identify (1) Twitter users who have posted tweets reporting that they took a beta-blocker during pregnancy and (2) their associated pregnancy outcomes, including miscarriage, stillbirth, birth defects, preterm birth, low birth weight, and neonatal intensive care unit (NICU) admission. We chose beta-blockers because cardiovascular disease is the leading cause of pregnancy-related deaths in the United States [6] and beta-blockers are the most common type of medication for treating cardiac conditions during pregnancy [7]. However, data on the safety of maternal beta-blocker exposure are inconsistent; some studies report associations low birth weight, preterm birth, perinatal mortality, or birth defects [8–16], while others do not [17–23].

## Methods

The Institutional Review Board (IRB) of the University of Pennsylvania reviewed this study and deemed it exempt human subjects research under Category (4) of Paragraph (b) of the US Code of Federal Regulations Title 45 Section 46.101 for publicly available data sources (45 CFR §46.101(b)(4)).

### Medication Intake

We searched for mentions of beta-blockers and their lexical variants (e.g., misspellings) [24] in 2.75 billion tweets posted by 415,690 users who have announced their pregnancy on Twitter [25]. Table 1 provides the beta-blocker keywords and their lexical variants. We manually reviewed the matching tweets to distinguish ones reporting that the user actually took the beta-blocker mentioned in the tweet [26]. To help determine if the user took the beta-blocker during pregnancy, we used the timestamp of the tweet reporting intake and drew upon an automated natural language processing (NLP) tool [27] that estimates the date of the user’s prenatal time period based on tweets that report the baby’s gestational age, due date, or date of birth. We also identified reports of taking a beta-blocker that occurred before or after pregnancy, assuming that, if there was no evidence in the tweet that the user stopped taking it before pregnancy or started taking it after pregnancy, the user may have been taking it during pregnancy. We excluded users for whom we could not estimate the date of their prenatal time period.

**Table 1.**
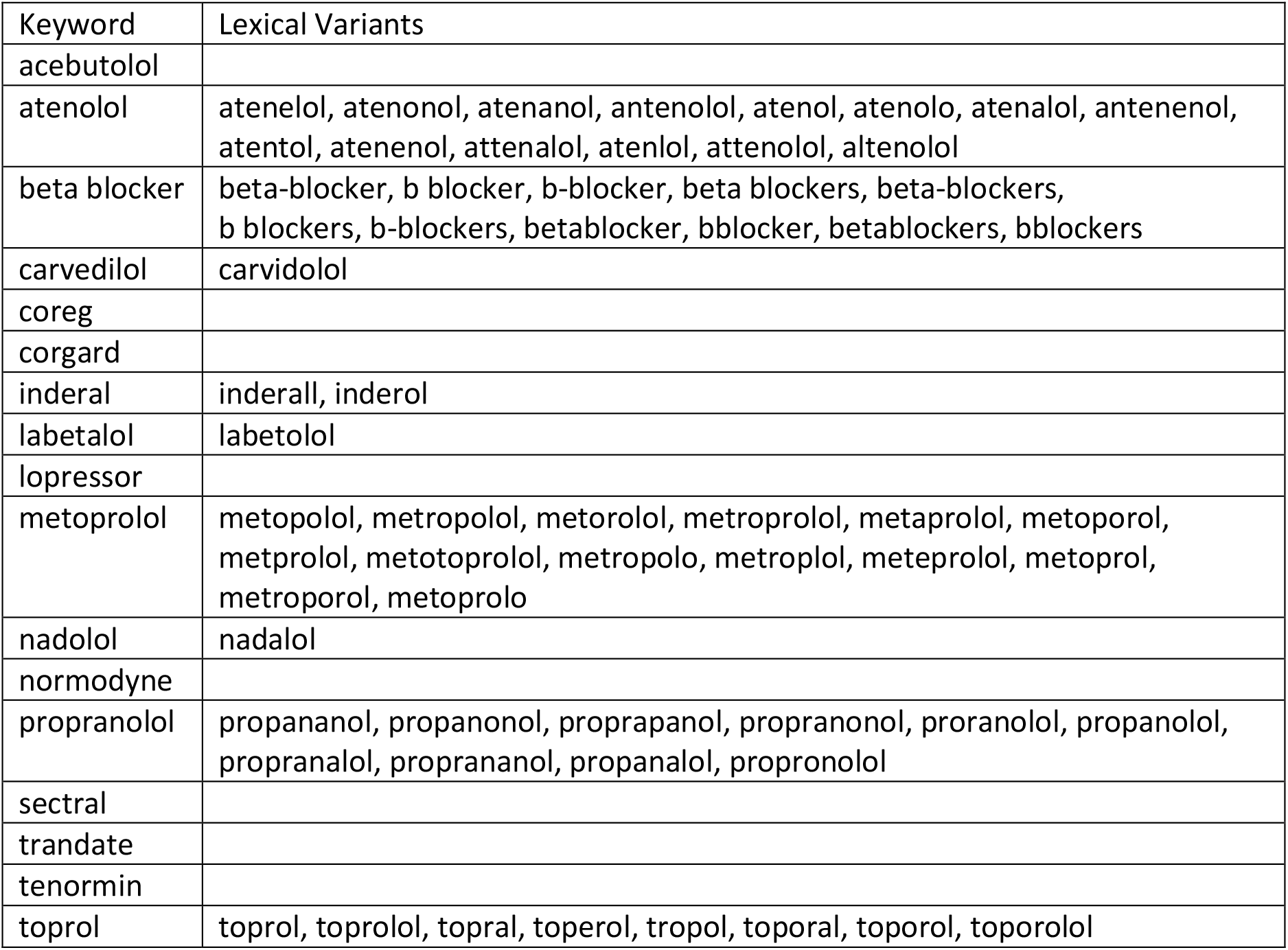
Keywords and their lexical variants used to search for tweets that mention beta-blockers.

### Pregnancy Outcomes

For users who posted tweets indicating that they took or may have taken the beta-blocker during pregnancy, we drew upon automated NLP tools [28,29] to help identify tweets that self-report an associated pregnancy outcome, including miscarriage, stillbirth, birth defects, preterm birth (<37 weeks gestation), low birth weight (<5 pounds and 8 ounces at delivery), or NICU admission. To assess a potential reporting bias (i.e., the lack of tweets self-reporting a pregnancy outcome may not represent the lack of that pregnancy outcome), we drew upon an automated NLP tool [30] that detects tweets reporting a gestational age ≥37 weeks (indicates the lack of miscarriage and preterm birth) or a birth weight ≥5 pounds and 8 ounces (indicates the lack of low birth weight, miscarriage, and stillbirth). If we did not automatically detect a tweet explicitly reporting a gestational age ≥37 weeks, we manually analyzed tweets posted during this time for evidence that the user was still pregnant.

### Covariates

Two important potential confounders when evaluating drug safety in pregnancy are maternal age and indication for use. To help identify maternal age, we deployed an automated NLP tool [31] that identifies tweets self-reporting the exact age of the user at the time the tweet was posted. Then, we used the date of the user’s prenatal time period to determine the user’s age during pregnancy. To identify an indication for use, we manually reviewed the tweets reporting intake of a beta-blocker posted by users who took or may have taken the beta-blocker during pregnancy.

## Results

Excluding retweets, we retrieved 5114 tweets, posted by 2339 users, that mention a beta-blocker, and manually identified 2332 (45.6%) tweets, posted by 1195 (51.1%) of the users, that self-reported taking the beta-blocker. We were able to estimate the date of the prenatal time period for 334 (27.9%) of the 1195 users. As some users’ collection of tweets span several years and include multiple pregnancies, we identified 356 pregnancies among these 334 users. Among these 356 pregnancies, we found evidence that a beta-blocker was or may have been taken in 257 (72.2%) of them: 58 (16.3%) pregnancies during which a beta-blocker was taken, and an additional 199 (55.9%) pregnancies during which a beta-blocker may have been taken (i.e., the report of taking a beta-blocker occurred before or after pregnancy, and there was no evidence in the tweet that the user stopped taking it before pregnancy or started taking it after pregnancy).

Table 2 presents examples of two users’ tweets that we used to determine their exposure to beta-blockers during pregnancy and their associated pregnancy outcomes. User 1 reported on January 25, 2020 that the baby’s due date was in 100 days, so our automated tool [27] estimated that pregnancy began on July 29, 2019 and would end on May 4, 2020. On April 16, 2020, User 1 explicitly reported taking Propranolol during pregnancy. User 1 reported that the baby was born premature on April 2, 2020—between 35 and 36 weeks gestation—and with a low birth weight of 4 pounds and 12 ounces, and was admitted to the NICU. User 2 reported being 37 weeks pregnant on June 1, 2020, so our automated tool [27] estimated that pregnancy began on September 16, 2019 and would end on June 22, 2020. Whereas User 1 explicitly reported taking a beta-blocker during pregnancy, for User 2, we used the timestamp of March 26, 2020 to infer that the intake was during pregnancy. User 2 reported on June 11, 2020 that the baby was born—between 38 and 39 weeks gestation—and weighed 7 pounds and 5 ounces at birth.

**Table 2.** Sample tweets used to determine exposure to beta-blockers during pregnancy and associated pregnancy outcomes.

| User | Tweet | Timestamp | Pregnancy start | Pregnancy end |
| --- | --- | --- | --- | --- |
| 1 | exactly 100 days til my due date! | 2020-01-25 | 2019-07-29 | 2020-05-04 |
|  | I was on Propranolol during my pregnancy and I had the CRAZIEST dreams I swear | 2020-04-16 |  |  |
|  | Officially introducing [name], born April 2nd, 2020. 4lbs 12oz, 18". She's in NICU due to being premature, but she's doing well! | 2020-04-03 |  |  |
| 2 | 5yo called me fat after I told 2.5yo I was too large to fit between their seats because of the baby. #37weekspregnant | 2020-06-01 | 2019-09-16 | 2020-06-22 |
|  | I saw the MFM and cardiologist last week. It was determined my cardiomyopathy is manageable and I was put on a beta blocker | 2020-03-26 |  |  |
|  | Introducing [name] 7lbs 5oz 20" long Csection went really well. We can't wait until the big boys get to meet him | 2020-06-11 |  |  |

We manually verified an adverse pregnancy outcome—preterm birth, NICU admission, low birth weight, birth defects, or miscarriage—for 38 (14.8%) of the 257 pregnancies during which a beta-blocker was or may have been taken. Table 3 presents the adverse pregnancy outcomes among these 257 pregnancies. We detected a gestational age ≥37 weeks for 198 (90.4%) of the 219 pregnancies for which we did not identify an adverse pregnancy outcome, and a birth weight ≥5 pounds and 8 ounces for 50 (22.8%) of these 219 pregnancies. We identified maternal age for 222 (86.4%) of the 257 pregnancies during which a beta-blocker was or may have been taken. Table 3 includes the mean age per adverse pregnancy outcome. We identified an indication for taking the beta-blocker for 197 (76.7%) of these 257 pregnancies—for example, tachycardia, hypertension, anxiety, and migraines.

**Table 3.**
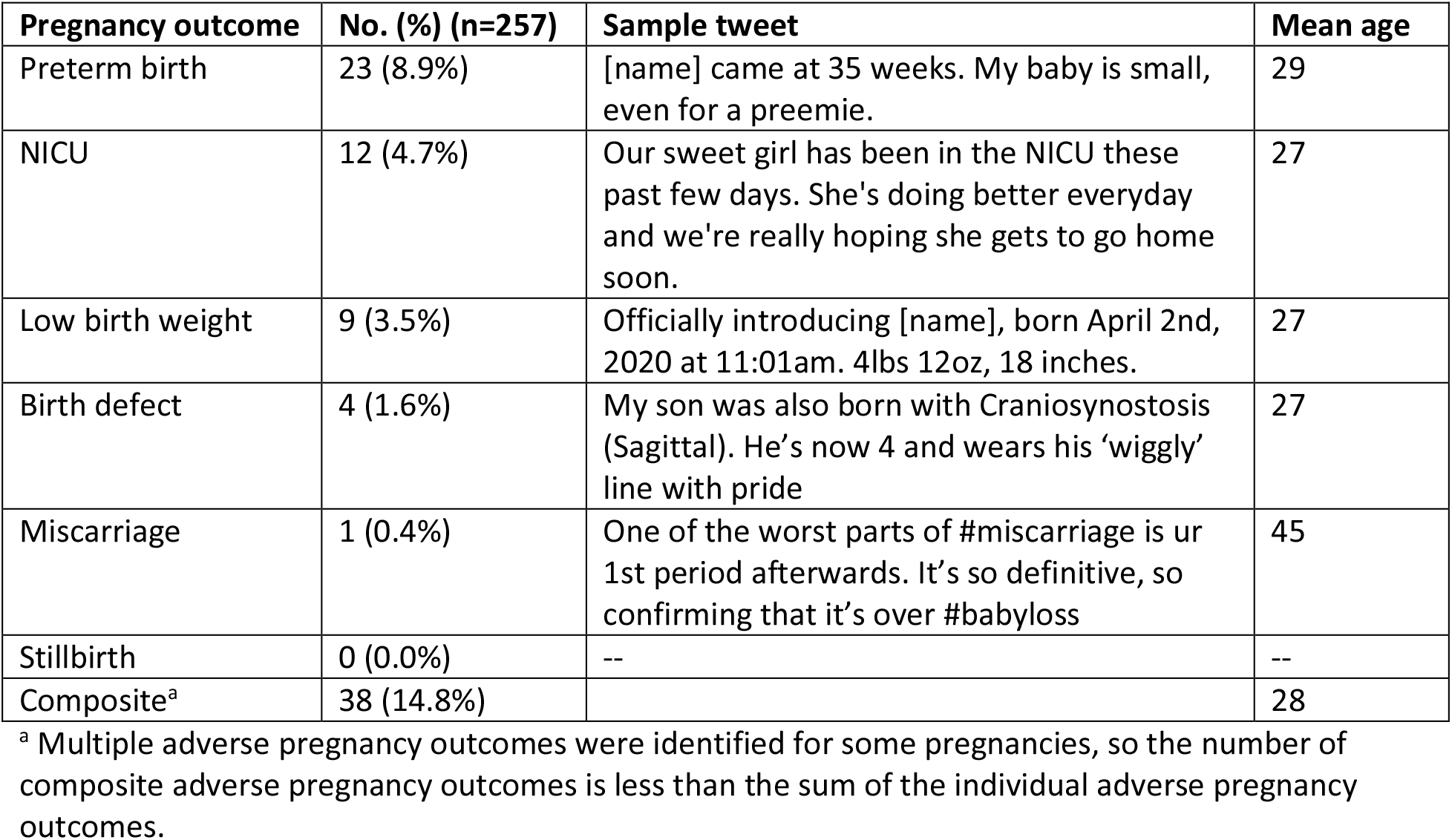
Self-reported adverse pregnancy outcomes for Twitter users who took or may have taken a beta-blocker during pregnancy.

| Pregnancy outcome | No. (%) (n=257) | Sample tweet | Mean age |
| --- | --- | --- | --- |
| Preterm birth | 23 (8.9%) | [name] came at 35 weeks. My baby is small, even for a preemie. | 29 |
| NICU | 12 (4.7%) | Our sweet girl has been in the NICU these past few days. She's doing better everyday and we're really hoping she gets to go home soon. | 27 |
| Low birth weight | 9 (3.5%) | Officially introducing [name], born April 2nd, 2020 at 11:01am. 4lbs 12oz, 18 inches. | 27 |
| Birth defect | 4 (1.6%) | My son was also born with Craniosynostosis (Sagittal). He's now 4 and wears his 'wiggly' line with pride | 27 |
| Miscarriage | 1 (0.4%) | One of the worst parts of #miscarriage is ur 1st period afterwards. It's so definitive, so confirming that it's over #babyloss | 45 |
| Stillbirth | 0 (0.0%) | -- | -- |
| Composite<sup>a</sup> | 38 (14.8%) |  | 28 |

<sup>a</sup> Multiple adverse pregnancy outcomes were identified for some pregnancies, so the number of composite adverse pregnancy outcomes is less than the sum of the individual adverse pregnancy outcomes.

## Discussion

Our ability to detect pregnancy outcomes for Twitter users who posted tweets reporting that they took or may have taken a beta-blocker during pregnancy suggests that Twitter can be a complementary resource for cohort studies of drug safety in pregnancy. Additionally, our ability to identify both the maternal age and indication for taking a beta-blocker for many of the users demonstrates that Twitter data would even allow such studies to account for the effect of these two important potential confounders. This study suggests that Twitter data may be particularly valuable for assessing associations with preterm birth, given both the volume of its reports on Twitter and our finding that preterm birth is largely unaffected by a potential reporting bias; that is, we detected a gestational age ≥37 weeks for 198 (90.4%) of the 219 pregnancies for which we did not identify an adverse pregnancy outcome. Low birth weight, however, may be affected by a potential reporting bias, given that we detected a birth weight ≥5 pounds and 8 ounces for only 50 (22.8%) of these 219 pregnancies. Although the rate of miscarriage in the United States is upward of more than 20% [32], our detection of miscarriage may be limited by a selection bias if users tend to announce their pregnancy on Twitter at a gestational age after which miscarriage infrequently occurs. Given our initial sample of 257 users, it is not surprising that we did not detect any reports of stillbirth, which has an incidence <1% in the United States [33]. Nonetheless, our prior work [29] demonstrates that users do report stillbirth outcomes on Twitter, and our identification of users announcing their pregnancy on Twitter continues to grow in real-time [25].

## Conclusions

Given the widespread use of medication during pregnancy and the insufficient data on fetal risks, Twitter can be a complementary resource for cohort study designs.

## Data Availability

All data produced in the present study are available upon reasonable request to the authors.

## Acknowledgments

AZK contributed to collecting the Twitter data, manually analyzing the Twitter data, and writing the manuscript. KO contributed to manually analyzing the Twitter data and editing the manuscript. LDL designed the study, including the selection of beta-blockers, pregnancy outcomes, and inclusion/exclusion criteria, and edited the manuscript. GGH conceptualized the use of Twitter data for studying medication exposure in pregnancy, guided the study, and edited the manuscript. The authors would also like to thank Ivan Flores for contributing to software applications, and Alexis Upshur for contributing to manually analyzing the Twitter data. This work was supported by the National Institutes of Health (NIH) National Library of Medicine (NLM; grant number R01LM011176).

